# A modest increase in ^11^C-PK11195-PET TSPO binding in depression is not associated with serum C-reactive protein or body mass index

**DOI:** 10.1101/2020.06.04.20099556

**Authors:** Julia J. Schubert, Mattia Veronese, Tim D. Fryer, Roido Manavaki, Manfred G. Kitzbichler, Maria A. Nettis, Valeria Mondelli, Carmine M. Pariante, Edward T. Bullmore, NIMA Consortium, Federico E Turkheimer

## Abstract

**BACKGROUND:** Immune mechanisms have been implicated in the pathogenesis of depression, and translocator-protein (TSPO) targeted positron emission tomography (PET) has been used to assess neuroinflammation in major depressive disorder. We aimed to: (i) test the prior hypothesis of significant case-control differences in TSPO binding in anterior cingulate (ACC), prefrontal (PFC) and insular (INS) cortical regions; and (ii) explore the relationship between cerebral TSPO binding and peripheral blood concentration of C-reactive protein (CRP).

**METHODS:** 51 depressed cases with Hamilton Depression Rating Scale score > 13 (median 17; IQR 16-22) and 25 healthy matched controls underwent dynamic brain ^11^C-PK11195 PET and peripheral blood immune marker characterisation. Depressed cases were divided into high CRP (>3mg/L;N=20) and low CRP (<3mg/L;N=31).

**RESULTS:** Across the three regions, TSPO binding was significantly increased in cases vs controls (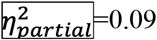; F(1,71)=6.97, *P*=0.01). which was not influenced by differences in body mass index (BMI). The case-control difference was greatest in ACC (d=0.49; t(74)=2.00, .*P*=0.03) and not significant in PFC or INS (d=0.27; d=0.36). Following CRP stratification, significantly higher TSPO binding was observed in low CRP depression compared to controls (d=0.53; t(54)=1.96, P=0.03). These effect sizes are comparable to prior MDD case-control TSPO PET data. No significant correlations were observed between TSPO and CRP measures.

**CONCLUSIONS:** Consistent with previous findings, there is a modest increase in TSPO binding in depressed cases compared to healthy controls. The lack of a significant correlation between brain TSPO binding and blood CRP concentration or BMI poses questions about the interactions between central and peripheral immune responses in the pathogenesis of depression.

## INTRODUCTION

There are a number of factors associated with major depressive disorder (MDD) and much recent research has focused on inflammation (1–6). Increases in markers of peripheral inflammation have previously been observed in individuals with MDD compared to healthy controls (HC) (6–9). Furthermore, inflammation and depression often occur together – “co-morbidly” – in the experience of patients with inflammatory diseases like rheumatoid arthritis or Crohn’s disease (10, 11), during treatment with pro-inflammatory cytokines (12, 13), and after experimental administration of a peripheral immune challenge, like typhoid vaccination (14, 15). An association has also been observed between inflammation and treatment-resistance to antidepressants (16, 17) and adjunctive antiinflammatory treatment has been shown to improve treatment efficacy of monoaminergic antidepressant drugs (18).

Previous studies in MDD have investigated the presence of neuroinflammation using positron emission tomography (PET) with radiotracers specific for 18-kDa translocator protein (TSPO) (1-6). TSPO is an outer mitochondrial membrane protein and elevations of TSPO expression have been consistently observed in microglial and macrophage populations during brain disease (19–22). Despite historically being used as a marker for microglial activity, TSPO is now known to be expressed in other cell types, including reactive astrocytes and endothelial cells (23–25). These cell types have also been shown to play a role in neuroinflammatory processes (26, 27). The association between TSPO expression in glial cell types involved in neuroinflammation enables the use of TSPO-specific ligand measurements to assess the presence and degree of neuroinflammation in neurological and psychiatric disease.

In studies of MDD cohorts, significantly higher TSPO binding compared to HC has been observed in the anterior cingulate by several groups (1–4), as well as higher binding in the frontal cortex (1–4, 6) and insula (2, 4). Relationships have also been observed between TSPO binding and medication status (3), length of time treated and untreated for MDD (2), and MDD disease duration (2). Holmes et al. (1) recently found that TSPO binding was significantly greater in MDD patients with suicidal thoughts compared to those without. A significant correlation between TSPO binding and depression severity scores in MDD has also been observed (4) and a reduction in TSPO binding has been observed in MDD patients undergoing cognitive behavioural therapy (6). However, other analyses have shown no difference in brain TSPO PET measures between MDD and HC (5) and did not observe links between TSPO PET measures and clinical scores (1, 5) or peripheral inflammatory markers (1, 4–6).

Here we collected clinical questionnaire data, TSPO PET brain scans, and peripheral blood immune markers from 51 depressed cases and 25 HC to better establish the relationship between peripheral and central inflammation in depression. ^11^C-PK11195, an isoquinoline carboxamide PET tracer specific for TSPO, was used with a dynamic PET acquisition to measure TSPO binding in the brain as a putative biomarker of central immune status. We tested the hypotheses: (i) that ^11^C- PK11195 binding measured in the three aforementioned regions-of-interest (ROIs), namely anterior cingulate, prefrontal cortex and insula, is significantly increased in depression; and (ii) that ^11^C-PK11195 binding is associated with blood concentration of C-reactive protein (CRP), as a biomarker of peripheral immune status, or body mass index (BMI). We also investigated whether TSPO binding is increased in treatment resistant or unmedicated depressed cases, and in depressed cases with suicidal thoughts compared to those without, in an attempt to replicate previous results.

## METHODS AND MATERIALS

### Participants

51 depressed cases (36/15 women/men; mean age: 36.2 ± 7.4 years) and 25 age- matched HC (14/11 women/men; mean age: 37.3 ± 7.8 years) were recruited from a network of clinical research sites in the United Kingdom as part of the Biomarkers in Depression study (BIODEP, NIMA consortium, https://www.neuroimmunology.org.uk/biodep/). Depressed individuals aged 25 to 50 (inclusive) on anti-depressant treatment with a total Hamilton Depression Rating Scale (HDRS)(28) score > 13 were included, as were untreated depressed subjects with a total HDRS > 17. Depressed cases were recruited and stratified into low/high CRP groups using a blood CRP concentration threshold of 3mg/L, resulting in 31 low CRP cases and 20 high CRP cases. During post-hoc analysis, depressed cases with HDRS > 13 despite more than 6 weeks treatment with two or more monoaminergic antidepressants were classified as treatment resistant and those not on treatment for at least 6 weeks prior to screening with HDRS > 17 were classified as untreated. Depressed cases with a score of 2 or higher on the ‘suicide’ item of HDRS were classified as having suicidal thoughts and those with a score of 0 on the ‘suicide’ item were classified as having no suicidal thoughts.

All cases and controls passed the following exclusion criteria: a lifetime history of other neurological disorders; active drug and/or alcohol abuse; participation in clinical drug trials within the previous year; concurrent medication or medical disorder that could compromise the interpretation of results; pregnancy or breastfeeding. Controls had no personal history of clinical depression requiring treatment and were mean age- matched with the cases. The study was approved by the NRES Committee East of England – Cambridge Central (REC reference: 15/EE/0092) and the UK Administration of Radioactive Substances Advisory Committee. All subjects gave written informed consent prior to data collection.

### Clinical Assessments

All subjects underwent an in-depth clinical evaluation that included the following psychiatric assessments: HDRS; Standard clinical interview for DSM (SCID); Beck’s Depression Inventory; Spielberger State-Trait Anxiety Rating Scale; Chalder Fatigue Scale; Snaith-Hamilton Pleasure Scale; and Perceived Stress Scale. Medical and family history was collected by a trained member of the research team. A venous blood sample was collected for assessing CRP level, as previously published (29). Venous blood was sampled from an antecubital vein between 08:00-10:00 h on the day of clinical assessment. Participants had fasted for 8 h, refrained from exercise for 72 h, and had been lying supine for 0.5 h prior to venepuncture. Blood was collected into a serum-separating tube, completely and gently inverted 10 times, allowed to coagulate for a minimum of 30 minutes and maximum of 60 minutes, and centrifuged at 1600 RCF for 15 minutes. 1mL of serum was then transferred with a pipette to a serum tube and sent on the day of collection to a central lab for high-sensitivity CRP assay via turbidimetric detection using a Beckman Coulter AU analyzer, with rabbit anti-CRP-antibodies coated on latex particles.

### TSPO PET data acquisition and analysis

Dynamic PET data acquisition was performed on a GE SIGNA PET/MR (GE Healthcare, Waukesha, USA) for 60 minutes after ^11^C-PK11195 injection (mean = 361 ± 53 MBq). Attenuation correction included the use of a multi-subject atlas method (30, 31) and improvements for the MRI brain coil component (32). Other data corrections (dead time, randoms, normalization, scatter, sensitivity, and decay) were as implemented on the scanner. Dynamic sinograms were reconstructed into 128 × 128 × 89 arrays (2.0 × 2.0 × 2.8 mm voxel size) using time-of-flight ordered subsets expectation maximization, with 6 iterations, 16 subsets and no smoothing. Examples of TSPO PET images from 10 to 60 minutes for one healthy control and one depressed case are shown in **Figure 1**. During PET data acquisition all subjects also had a volumetric, high resolution T1-weighted brain MRI (BRAVO), which was used for PET data processing.

**Table 1.** Demographic and clinical characteristics for depressed cases and healthy controls

| Variable | Depressed cases<br>(n = 51) | Healthy controls<br>(n = 25) | P Value |
| --- | --- | --- | --- |
| Age, mean (SD), y | 36.2 (7.3) | 37.3 (7.8) | 0.561 |
| Male, No. (%) | 15 (29) | 11 (44) | NA |
| Weight, mean (SD), kg | 80.3 (14.4) | 73.7 (15.1) | 0.102 |
| BMI, mean (SD), kg/m<sup>2</sup> |  |  |  |
| All | 27.2 (4.0) | 24.2 (4.8) | 0.001* |
| High CRP | 30.0 (3.8) | NA | NA |
| Low CRP | 26.2 (3.4) | NA | NA |
| CRP, No., mean (SD), mg/L |  |  |  |
| All | 51, 2.9 (2.8) | 25, 1.1 (0.9) | <0.001* |
| High CRP | 20, 5.6 (2.6) | NA | NA |
| Low CRP | 31, 1.1 (0.7) | NA | NA |
| Medication status, No. (%) |  |  |  |
| Untreated | 9 (18) | NA | NA |
| Taking medication | 42 (82) | NA | NA |
| Treatment resistant | 33 | NA | NA |
| HDRS, mean (SD) | 18.5 (3.7) | 0.6 (0.9) | <0.001* |
| Dose of radioactive isotope injected,<br>mean (SD), MBq | 360.3 (53.2) | 376.2 (44.8) | 0.138 |
| InjectedMass, mean (SD), µg | 3.0 (1.6) | 3.4 (1.8) | 0.486 |
| specificActivity, mean (SD), GBq/µmol | 50.80 (21.33) | 50.56 (25.94) | 0.711 |
| Total Motion, mean (SD), mm | 7.76 (3.75) | 7.27 (3.39) | 0.562 |
| Max InterFrame Motion, mean (SD), mm | 1.62 (0.90) | 1.45 (0.68) | 0.532 |
| Scan Time, hh:mm:ss | 15:14:44 | 14:58:05 | 0.761 |

Abbreviations: NA, not applicable

**Figure 1.**
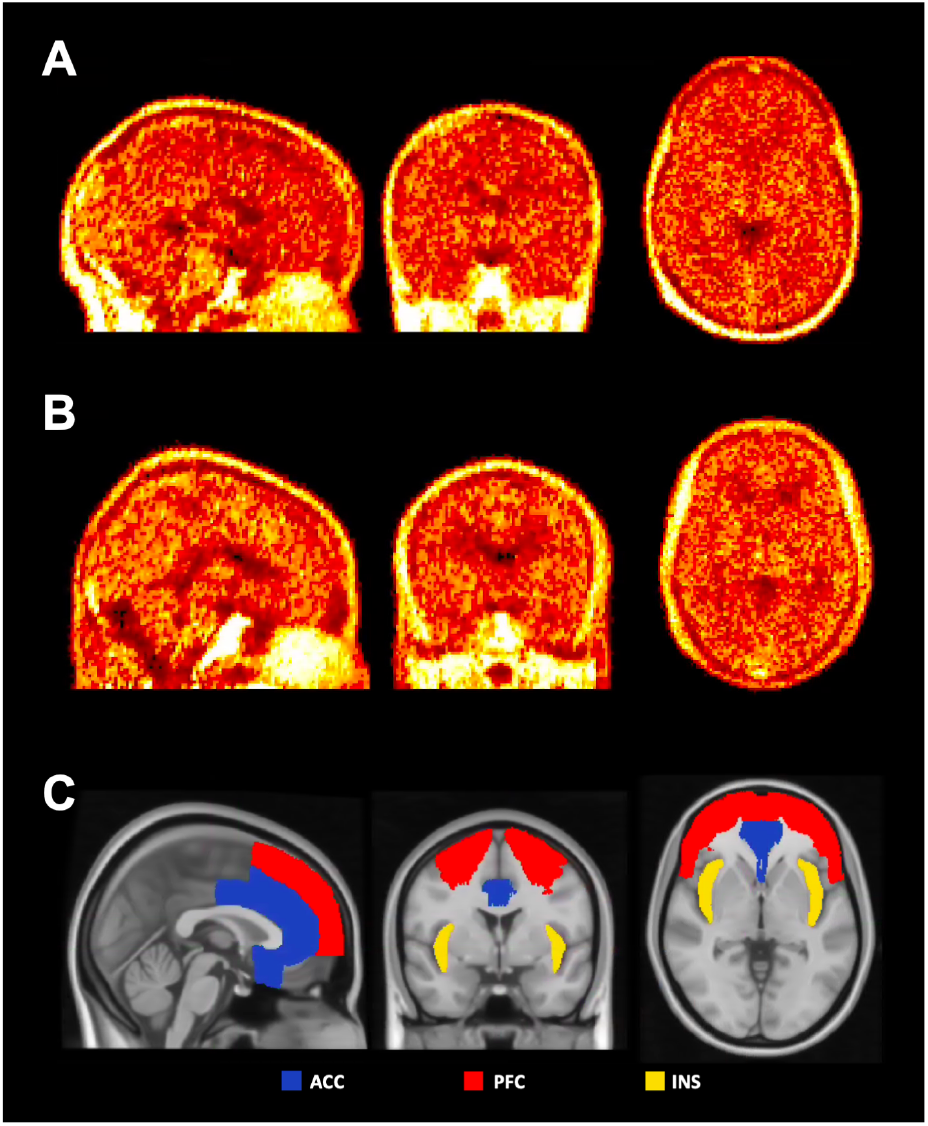
Illustrative examples of ^11^C-PK11195 PET images from 10 to 60 minutes for one healthy control (A) and one depressed case (B). C:Anterior cingulate cortex (ACC), prefrontal cortex (PFC), and insular cortex (INS) regions overlaid on MNI152 brain template.

Brain extraction, tissue segmentation, alignment of the MRI and PET data, and motion correction were performed using MIAKAT™ (version 4.2.6) software. MIAKAT™ is implemented in MATLAB (version R2015b; The MathWorks, Inc., Natick, Mass.) and uses tools from SPM12 and FSL (version 5.0.9). Data quality control was performed through visual inspection of the outputs of the MRI and PET processing steps. Experimental variables including injected activity, total motion during PET, and maximum interframe motion during PET were recorded for all subjects (**Table 1**). The CIC v2.0 neuroanatomical atlas was coregistered to the image space of each subject and used to extract time-activity curves from a subset of 24 ROIs based on their relevance to MDD. A simplified reference tissue model using a supervised clustering reference region approach (33) was used to quantify ^11^C-PK11195 binding as relative binding potential (BP_ND_) in three primary bilateral ROIs (anterior cingulate (ACC), prefrontal cortex (PFC), and insula (INS)), which are shown in **Figure 1C**. Example time activity curves for ACC of one healthy control and one depressed case are shown in **Figure 2A** and **2B**, respectively. The supervised approach uses a predetermined set of kinetic tissue classes to identify voxels with kinetic behavior closest to that of healthy gray matter. The time-course of the activity averaged over the identified voxels, weighted by the gray matter kinetic class scaling coefficient per voxel, is used as the reference input function. Use of a supervised cluster analysis technique for extracting reference tissue has previously been cross-validated and shown to be a reliable method for quantifying brain ^11^C-PK11195 (34). The three ROIs were selected based on previous findings (1, 4), which were originally motivated by their role in mood regulation (35, 36), together with previous suggestions of the involvement of the ACC in the link between inflammation and depression (14, 37–41). The PFC region used in this analysis is the aggregate of the medial and dorsolateral frontal cortex regions from the CIC v2.0 neuroanatomical atlas.

**Figure 2.**
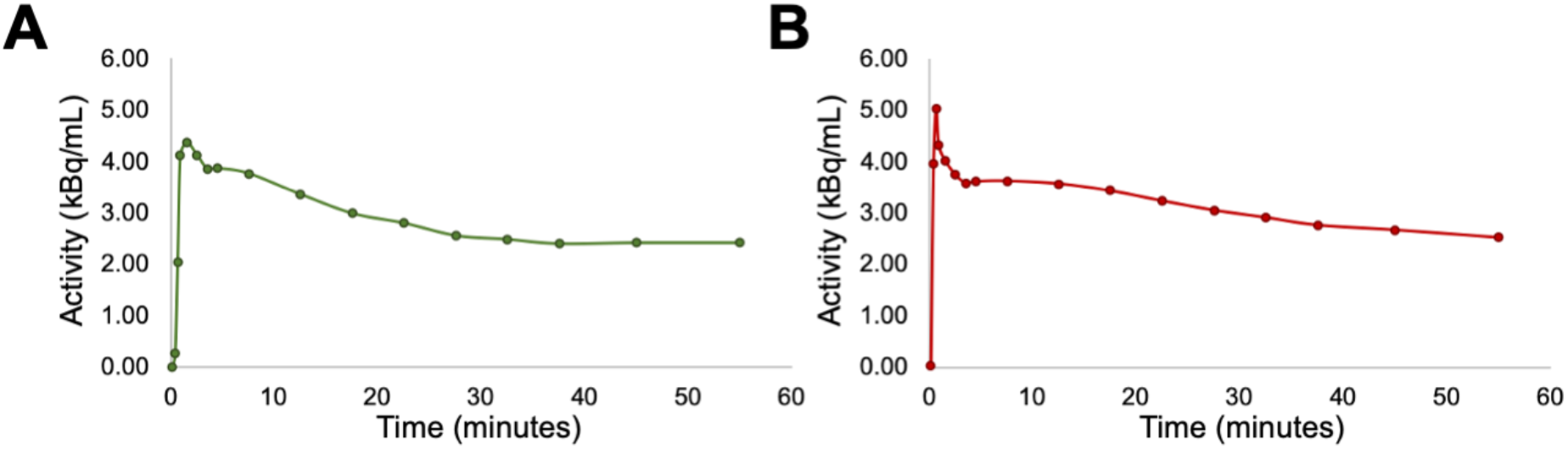
Example time activity curve for ACC of a healthy control subject (A) and depressed case (B).

### Statistical Analysis

SPSS (version 24.0, Chicago, IL) was used to perform all statistical analyses. Normality of the data was tested using Shapiro-Wilk’s W test. BP_ND_ across the ACC, PFC, and INS regions was investigated using analysis of variance (ANOVA) to test for case-control differences, while covarying for age, sex, and BMI. Group differences in experimental variables (subject age, body weight, injected activity, total motion, maximum interframe motion), and BP_ND_ in individual ROIs, were further investigated using the independent-samples Mann-Whitney U-test, and one-tailed independent samples t-test, respectively. Group differences were evaluated with 5% probability of type 1 error (α=0.05). The relationships between CRP concentration and BP_ND_, and between psychiatric assessment scores and BP_ND_ in the three primary ROIs, were assessed using Spearman’s correlation. Estimated sample sizes required to achieve significant group differences in PFC and INS regions were calculated given the observed effect sizes, with α=0.10 and β=0.20. Nominal p-values are reported without correction for multiple comparisons.

## RESULTS

### Demographic and clinical characteristics

Demographic, clinical and PET scan details for healthy controls and depressed cases are given in **Table 1**. There are significant differences in BMI (*P*=0.001), serum CRP concentration (*P*<0.001), and HDRS (*P*<0.001) between depressed cases and healthy controls. No other significant differences between healthy controls and depressed cases were observed for PET imaging parameters.

### TSPO tracer binding comparisons between HC and depression

ANOVA revealed a significant case-control difference in BP_ND_ across the three primary ROIs (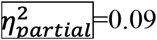; F(1,71)=6.97, *P*=0.01) **(Figure 3A)**. A significantly higher average BP_ND_ was observed in the ACC in depressed cases (mean = 0.17, SD = 0.04) compared to healthy controls (mean = 0.15, SD = 0.05) (d=0.49; t(74)=2.00, *P*=0.03) which remained significant when covaried for age, sex, and BMI (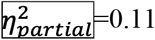; F(1,71)=9.07, *P*=0.004). There were no other significant differences in BP_ND_ between groups in the PFC (d=0.27; t(74)=1.11, *P*=0.13) or INS regions (d=0.36; t(74)=1.48, *P*=0.07), at nominal 5% probability of type 1 error. These results are shown in the context of previously reported case-control differences in ^11^C-PKl 1195 binding in a forest plot in **Figure 3B**. Estimated sample sizes required to achieve significant differences with the observed effect sizes are N=252 for PFC and N=116 for INS, assuming equal number of subjects in case and control groups.

**Figure 3.**
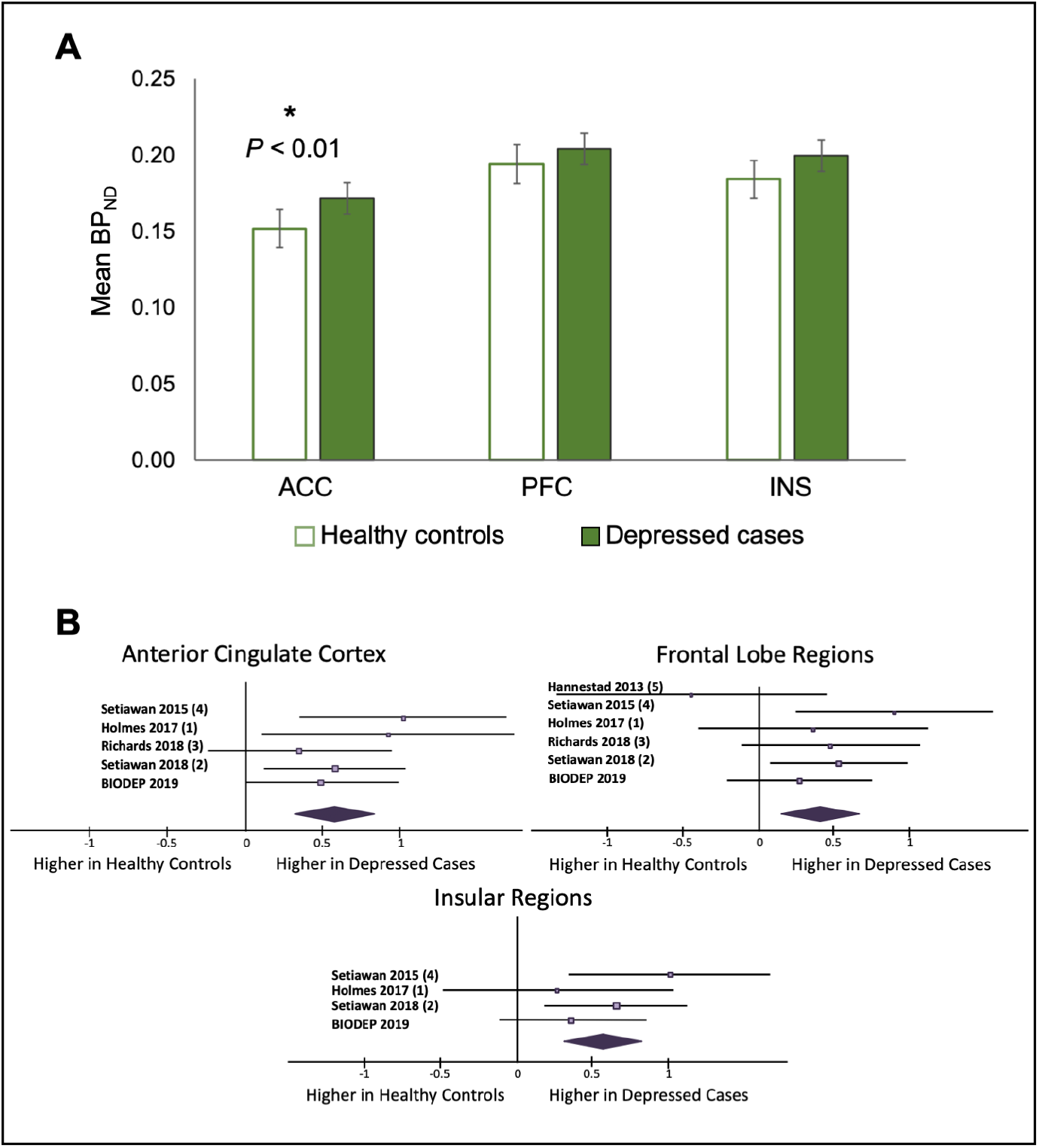
**A**. Case-control differences in mean ^11^C-PK11195 binding potential measurements in anterior cingulate cortex (ACC), prefrontal cortex (PFC), and insular cortex (INS) regions between healthy controls and depressed cases. Error bars represent standard error. The analyses are corrected for age, sex, and BMI. Significant differences (*P*<0.05) are indicated by an asterisk. **B**. Forest plot summarising the current results in the context of previous TSPO PET results from case-control studies of depression in the anterior cingulate cortex, frontal lobe regions, and insular regions.

### Relationships between TSPO tracer binding and CRP

A significantly higher average BP_ND_ was observed in the ACC in *low* CRP depressed cases compared to healthy controls (d=0.53; t(54)=1.96, *P*=0.03) before correcting for age, sex, and BMI. Significantly higher average BP_ND_ was observed in ACC in high CRP depressed cases compared to healthy controls (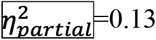; F(1,40)=5.74, *P=0.02)*, in ACC in low CRP depressed cases compared to healthy controls (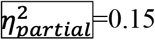; F(1,51)=8.82, *P*=0.01) and in INS in high CRP depressed cases compared to healthy controls ((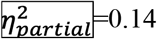; F(1,40)=6.68, *P*=0.01) after correcting for age, sex, and BMI **(Figure 4)**. No other significant differences were observed when the depressed group was stratified by CRP concentration. No significant correlations were observed between BP_ND_ and CRP, HDRS, or BMI in the ROIs **(Figure 5)**.

**Figure 4.**
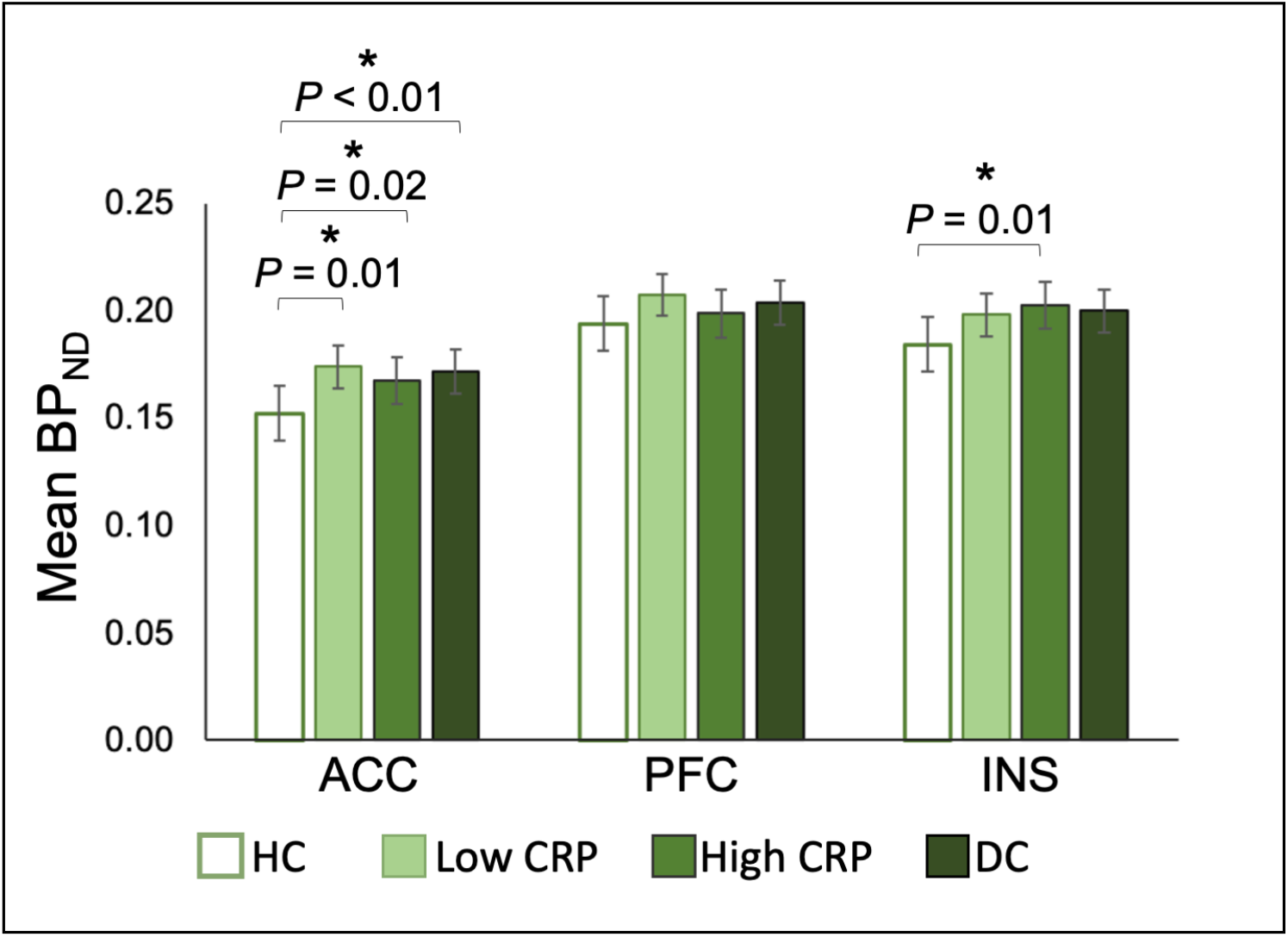
Case-control differences in mean ^11^C-PKlf 195 binding potential measurements in anterior cingulate cortex (ACC), prefrontal cortex (PFC), and insular cortex (fNS) regions between healthy controls (HC), low CRP depression, high CRP depression, and all depressed cases (DC). Error bars represent standard error. Analyses are corrected for age, sex, and BMI Significant differences (*P*<0.05) are indicated by an asterisk.

**Figure 5.**
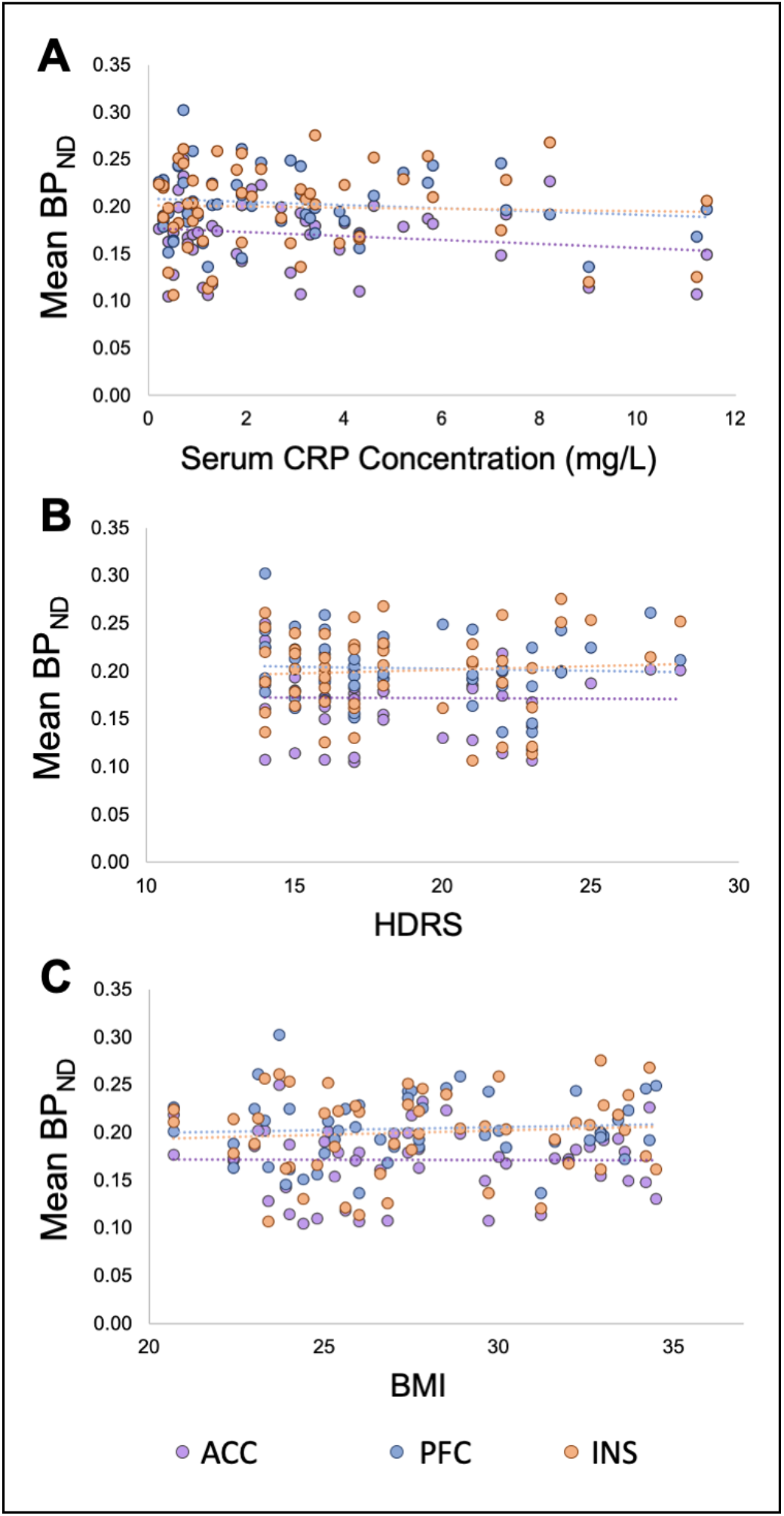
Scatterplot of ^11^C-PK11195 binding potential measurements (y-axis) for anterior cingulate cortex (ACC), prefrontal cortex (PFC), and insular cortex (INS) regions versus serum C-reactive protein (CRP) concentration (mg/L) (A), Hamilton depression rating scale (HDRS) (B), and body mass index (BMI) (C) in depressed cases.

#### Post-hoc investigations of TSPO tracer binding and clinical variables

No significant correlations were observed between BP_ND_ and HDRS scores, as the primary score to investigate disease severity in depression **(Figure 5B)**. Further exploratory analysis showed no significant correlations between BP_ND_ and other psychiatric assessment scores including Beck’s Depression Inventory, Spielberger State-Trait Anxiety Rating Scale, Chalder Fatigue Scale, Snaith-Hamilton Pleasure Scale, and Perceived Stress Scale. Differently from previous studies, no significant differences were observed between HC and treatment resistant depression, between HC and untreated depression **(Figure S1A)**, between HC and depressed cases with suicidal thoughts, or between depressed cases with and without suicidal thoughts **(Figure S1B)**.

A complete report of the primary statistical analysis results is included in Supplementary **Tables S1-S3**. Correlation matrices showing the relationship between regional BP_ND_, CRP, HDRS, and BMI in depressed cases and healthy controls are included in Supplementary **Tables S4** and **S5**, respectively.

## DISCUSSION

This study successfully replicates previous results of increased TSPO binding in depressed cases compared to healthy controls, in the largest to-date sample of TSPO PET data for investigating neuroinflammation together with peripheral inflammation in depression. Consistent with the study hypothesis, we observed an increase in TSPO binding in depressed cases compared to healthy controls across the three primary ROIs (ACC, PFC and INS). However, we did not observe any significant correlation between brain TSPO PET measures and serum CRP concentration or BMI or clinical scores. We did not replicate previous findings of increased TSPO binding in untreated cases compared to controls (3), or in cases with suicidal thoughts compared to cases without suicidal ideation (1). We observed greater variability in BP_ND_ values in all three primary ROIs in the HC group compared to depressed cases.

TSPO PET studies may be difficult to compare due to use of different radiotracers and quantification measures (42). Even so, recent studies (1–4) along with ours indicate that there is increased TSPO binding in depressed cases compared to healthy controls, where the meta-analytic effect size is medium on average over all studies (overall Hedge’s g for ACC:0.60, PFC: 0.41, INS:0.57). Generally these results provide robust support for the presence of somewhat greater central nervous system inflammation in depressed cases compared to HC. Our result of no association between CRP and TSPO binding is also consistent with recent literature (1, 3, 4, 6) and suggests that, although an association has been observed between increased peripheral inflammation and MDD (43), CRP as a marker for peripheral inflammation is not associated with central inflammation putatively measured with TSPO PET. BMI is one of the main contributing factors to chronic low-grade inflammation (44) and has been identified as a confounding factor in TSPO PET studies (46). However, depression is associated with peripheral inflammation even after adjusting for BMI (29, 49, 53). The lack of association between BMI and PET signal furher confirms that peripherally-produced inflammation does not translate directly into central inflammation. Consistent findings of weak central inflammation in depressed cases compared to healthy controls, and the lack of a direct association between measures of central and peripheral inflammation, do not seem to match the associations between peripheral and central inflammation observed in animal models (45). Previous findings have posed the model of depression involving direct induction of central immune activation by peripheral cytokines, which in turn could lead to a biochemical cascade that eventually leads to depressive symptoms (47, 48). However, if this model was to apply in our cohort, we would expect less variability in the TSPO result of the depressed group and to observe an association between central (TSPO PET) and peripheral (CRP) inflammatory measures, which we do not. In fact most animal models of inflammation-induced sickness behaviour are characterized by a leaky blood-brain barrier (45) and MDD has only been observed to exhibit a leaky blood-brain barrier in a few cases (50), suggesting that these animal models may be inappropriate for representing the general relationship between the central nervous system and the periphery in MDD. TSPO PET results investigating the link between central and peripheral inflammation by peripheral immune challenge in humans are also contradictory. One study showed that injection of Escherichia coli lipopolysaccharide significantly increased measures of brain TSPO (51). Another showed that immune challenge with interferon alpha exhibited no change in brain TSPO (52) and both studies showed no link between peripheral inflammation, measured by serum CRP, and brain TSPO (51, 52).

This does not imply that peripheral inflammation and depression are otherwise unrelated. Increased peripheral (53, 54) and central (1, 3, 4, 6) inflammatatory markers have been separately observed in MDD cases that do not present with inflammatory comorbities, which suggests that both peripheral and central inflammation may play a part in the development or progression of MDD. However the mechanism linking peripheral cytokine activity and central immunity is still unclear; our results along with those from previous groups suggest that this complex relationship warrants further investigation in more realistic settings. Interestingly, some of our findings became more significant or apparent when controlling the statistical analyses for BMI. It is well acknowledged that metabolic and immune system are strongly integrated and that higher BMI is associated with higher levels of peripheral inflammation, including higher CRP levels (58). The finding of significantly higher TSPO binding in depressed cases even after correcting for BMI again suggests that a peripheral metabolic-immune dysfunction per se is not sufficient to induce increase in TSPO expression or development of depression and other factors, such as BBB permeability, may be relevant for a better understanding of the link between peripheral metabolic-immune dysfuntion and depression.

### Limitations

This study has some limitations. The greater variability of the BP_ND_ values for all regions in the HC compared to depressed group presented here is surprising. A possible explanation for this is the higher proportion of males in the HC group, who have previously shown to have higher variance in BP_ND_. Recruitment of truly healthy controls is challenging but questionnaires about current and past medical problems were analyzed for all subjects to ensure that cases and controls with potentially confounding medical co-morbities were excluded. The TSPO PET signal may be difficult to interpret at the cellular level because TSPO is not specific to microglia. TSPO is also upregulated in other cell types during disease (22, 24, 55). Other cell types that express TSPO include macrophages, astrocytes, epithelial and vascular endothelial cells, although they all have a role in CNS immunity (55, 56). In any case, more recent evidence has also shown that microglial expansion does correspond to TSPO binding in tissue during disease (24). The future availability of more specific markers for microglial activation and neuroinflammation (57, 59–65), to be used in place of or in parallel to TSPO ligands, will substantially help the interpretation of future results in the area of neuroinflammation and depression.

### Conclusions

We have contributed to the growing body of work (1–4, 6) that continues to support a relationship between neuroinflammation and depression. However, the effect size of case-control differences in the TSPO PET signal is small and is not correlated with peripheral CRP concentrations suggesting the scope for future, more mechanistic studies that will require novel PET radioligands with specific brain immune targets.

## Data Availability

The datasets generated during and/or analysed during the current study are available from the corresponding author on reasonable request, and upon approval from the research consortia.

## ACKNOWLEDGEMENTS AND DISCLOSURES

The BIODEP study was sponsored by the Cambridgeshire and Peterborough NHS Foundation Trust and the University of Cambridge, and funded by a strategic award from the Wellcome Trust (104025) in partnership with Janssen, GlaxoSmithKline, Lundbeck and Pfizer. Recruitment of participants was supported by the National Institute of Health Research (NIHR) Clinical Research Network: Kent, Surrey and Sussex & Eastern. Additional funding was provided by the National Institute for Health Research (NIHR) Biomedical Research Centre at South London and Maudsley NHS Foundation Trust and King’s College London, and by the NIHR Cambridge Biomedical Research Centre (Mental Health). ETB and CMP are supported by a Senior Investigator award from the NIHR. Study data were collected and managed using REDCap electronic data capture tools hosted at the University of Cambridge (66).

We would like to gratefully thank all study participants, research teams and laboratory staff, without whom this research would not have been possible. All members of the NIMA Consortium at the time of data collection are thanked and acknowledged in the Appendix.

